# Venezuelan Equine Encephalitis Virus, Amazonas, Brazil, 2025

**DOI:** 10.1101/2025.08.19.25333804

**Authors:** Franklin Simões de Santana Filho, Marcelo Ferreira dos Santos, Victor Costa de Souza, Valdinete Alves do Nascimento, Maria do Perpétuo Socorro Lopes dos Santos, Gian Pierre Hernandez Medina, Klaus Estivens Lima Salazar, Jhamylee Evellyn Sanchez Mendez, Elizabeth Andreia da Silva Brito Fajardo, Luis Castelo Branco de Oliveira, David Siqueira da Costa, Tyziane Machella Oliveira Torres, Ana Cláudia Magalhães Salvador, Syssi Helen Albuquerque do Nascimento, Bruna Oliveira da Silva, Karina Lopes Oliveira, Herton Augusto Pinheiro Dantas, Matilde Mejía, Fernanda Nascimento, Dejanane Silva, Maria Julia Pessoa Brandão, Ana Beatriz Vieira Muniz, Tatiana Amaral Pires de Almeida, Marco Aurélio Almeida de Oliveira, Luciana Mara Fé Gonçalves, Tatyana Costa Amorim Ramos, Flor Ernestina Martinez-Espinosa, André Reynaldo Santos Périssé, Deusilene Souza Vieira Dall’Acqua, Ana Maria Bispo de Filippis, Gonzalo Bello, Martha Cecília Suárez-Mutis, Felipe Gomes Naveca

**Author notes:** Corresponding author: Felipe Gomes Naveca,Address: Instituto Leônidas e Maria Deane (ILMD), Fiocruz Amazônia. 476 Teresina Street, Adrianópolis, Manaus - Amazonas, 69057-070, Brazil & Instituto Oswaldo Cruz. 4365 Brazil Avenue, Rio de Janeiro - Rio de Janeiro, 21040-360, Brazil.

## Abstract

We report the first detection of Venezuelan Equine Encephalitis Virus (VEEV) in residents of Tabatinga, Amazonas, Brazil, a tri-border region with Colombia and Peru. Brazilian and Peruvian sequences clustered together, suggesting potential cross-border transmission. Our findings identify VEEV as an underrecognized etiology of acute febrile illness in the Brazilian Amazon.

## The Study

The tri-border Amazonian region of Brazil, Colombia, and Peru represents a complex ecotone with high biodiversity and extensive human mobility, creating optimal conditions for disease emergence and cross-border transmission. Acute febrile illness (AFI) is a primary cause of healthcare-seeking behavior in this region; however, its underlying etiology often remains underdiagnosed (1). This diagnostic challenge is further complicated by symptom overlap with endemic diseases, such as malaria and arboviruses, which can obscure other circulating pathogens (2,3). Implementing laboratory surveillance with expanded molecular testing panels could be a critical strategy in endemic areas with undifferentiated fever (4).

Venezuelan equine encephalitis virus (VEEV) is an arbovirus that belongs to the genus *Alphavirus* (family *Togaviridae*). According to the International Committee on Taxonomy of Viruses, it is the prototype of the *Alphavirus venezuelan* species, a complex that includes multiple subtypes with distinct ecological niches and pathogenic potential (https://ictv.global/sites/default/files/VMR/VMR_MSL40.v1.20250307.xlsx). It persists in nature through two primary transmission cycles: an enzootic cycle involving rodents as primary hosts and *Culex (Melanoconion) spp*. mosquitoes as vectors for virus subtypes ID–IF and II–VI and an epizootic cycle where equines act as amplification hosts for virus subtypes IAB and IC, which are primarily transmitted by *Aedes* and *Psorophora spp*. vectors. Recognized as a human pathogen since the late 1950s, VEEV typically causes an undifferentiated AFI, though a minority of cases can progress to severe, sometimes fatal, neuroinvasive disease (5).

Our study, as part of the “Front Fever” project, is a cross-sectional investigation initiated in July 2024. We enrolled adult patients with AFI (axillary temperature ≥38.0°C) in Tabatinga (Brazil), in the tri-border region with Colombia and Peru (**Figure 1**). Blood and oronasal swab samples were collected for molecular analysis after excluding cases with evident infectious foci, including malaria. We employed a two-tiered diagnostic workflow to ensure thorough investigation of each case. Accordingly, our team performed real-time RT-PCR screening for the most common regional arboviruses and respiratory viruses, followed by analysis of the undiagnosed samples applying an expanded real-time RT-PCR panel. Although the broader “Front Fever” project investigates multiple etiologies, this manuscript focused on the arbovirus diagnostic workflow that led to the detection of VEEV. The Research Ethics Committee of the Federal University of Amazonas approved this study (protocol number 123.456 / CAAE: 78571324.0.0.0000.5020), and each patient signed informed consent.

**Figure 1.**
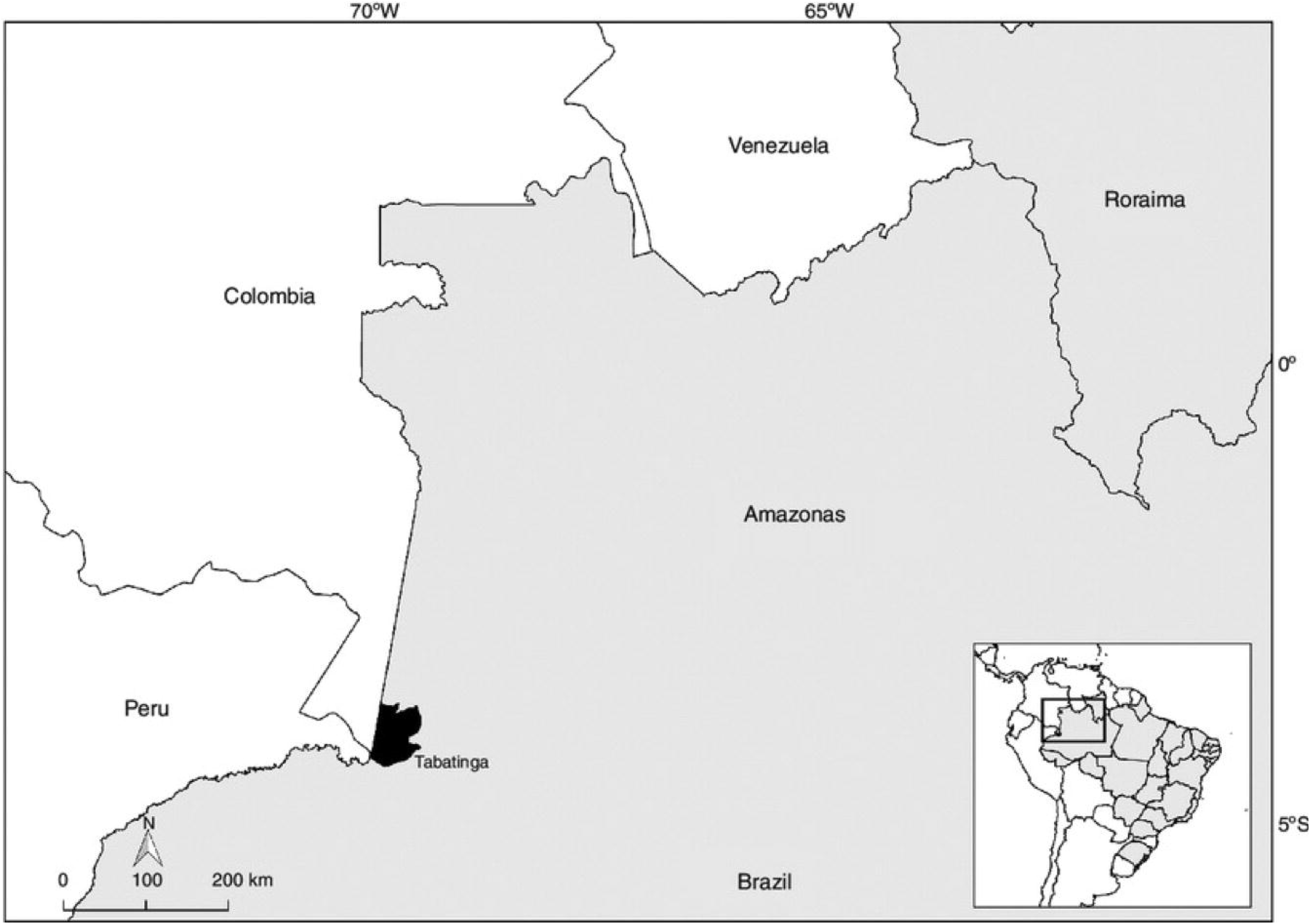
Study site and location of VEEV cases in Tabatinga, Amazonas, Brazil. Map of the Brazilian Amazon, highlighting the municipality of Tabatinga in the tri-border region with Colombia and Peru.

In July 2025, we detected and reported three human infections with VEEV to the health authorities of the municipality of Tabatinga, Amazonas State, and the Brazilian Ministry of Health. Initial real-time RT-PCR screening for primary arboviruses (Dengue serotypes 1–4, Zika, and Chikungunya viruses) was performed locally at the Frontier Laboratory of Public Health (LAFRON) using the commercial kit Biomol ZDC (IBMP, Curitiba, Brazil), followed by in-house testing for Mayaro and Oropouche viruses (6). A total of 80 undiagnosed plasma samples were subsequently referred to Instituto Leônidas e Maria Deane (ILMD - Fiocruz Amazônia) for further etiological investigation using a comprehensive in-house arbovirus diagnostic panel (Yellow Fever, West Nile, St. Louis encephalitis, Eastern and Western equine encephalitis, VEEV, and O’Nyong-Nyong viruses).

This exhaustive molecular investigation identified three human infections with VEEV. The cycle threshold (Ct) values from VEEV-positive real-time RT-PCR tests were 35.7, 36.5, and 38.4, respectively (**Table 1 and Supplemental Figure 1A**), indicating low viral loads or potential issues with mismatched primers or probes. All three patients were residents of Tabatinga urban area and presented with a nonspecific AFI with systemic symptoms such as headache, myalgia, and arthralgia, with no signs of neuroinvasive disease (**Table 1**). The interval between the onset of symptoms and their visit ranged from three to five days. Notably, two of the three patients presented with leukopenia and thrombocytopenia, laboratory findings that complicate clinical differentiation from other viral infections, such as dengue. At the scheduled follow-up on day 28, all three patients had fully recovered without any sequelae. No human or equine encephalitis outbreaks were officially reported in this region during the study period.

**Table 1.** Summary of VEEV infection cases in Tabatinga, Brazil, 2025.

| Characteristic | Case 1 (ID 051) | Case 2 (ID 055) | Case 3 (ID 063) |
| --- | --- | --- | --- |
| Collection Date | February 2025 | March 2025 | March 2025 |
| Age range / Sex | 50-59 years / M | 50-59 years / F | 30-39 years / M |
| Occupation | Farmer | Homemaker | Healthcare Professional |
| Residence | Urban Area | Urban Area | Urban Area |
| Relevant Exposure | Work in the forest | "Carapanã" mosquito bites* | "Carapanã" mosquito bites* / Equine proximity |
| Laboratory Findings |  |  |  |
| Leukocytes | 4,000/mm<sup>3</sup> | 2,400/mm<sup>3</sup><br>(Leukopenia) | 3,000/mm<sup>3</sup><br>(Leukopenia) |
| Platelets | 182,000/mm<sup>3</sup> | 130,000/mm<sup>3</sup><br>(Thrombocytopenia) | 133,000/mm<sup>3</sup><br>(Thrombocytopenia) |
| CRP | 3.0 mg/L | 11.0 mg/L | 10.0 mg/L |
| VEEV RT-PCR (Ct) | 35.7 | 36.5 | 38.4 |

\*The term “carapanã” is a regional Brazilian designation for mosquitoes, particularly those belonging to the family *Culicidae*, including species of the genus *Anopheles*. CRP: C-reactive protein. Ct: cycle threshold value.

For etiological confirmation and characterization, samples positive for VEEV underwent conventional RT-PCR targeting the nsP1 gene, followed by capillary sequencing and maximum-likelihood (ML) phylogenetic analysis. Viral RNAs were subjected to conventional RT-PCR amplification of the nsP1 protein-coding region using the M2W and cM3W primers **(7)**, resulting in an amplicon of 434 bp (**Supplemental Figure 1B**). Subsequent capillary sequencing of this product yielded a 312bp consensus sequence. BLAST analysis initially confirmed that the sequence was a VEEV, with all top hits exhibiting significant E-values (down to 9E-157). Thereafter, we designed other primers to amplify and sequence a larger fragment of 854bp (807bp after trimming primers) from sample Tabatinga 055 (**Supplemental Figure 1C**), which was then used for a new ML phylogenetic reconstruction. This analysis revealed that Tabatinga 055 sequence clustered within a monophyletic clade alongside other VEEV subtype ID sequences of Peruvian origin (**Figure 2**). Currently, health authorities across the region are instituting surveillance efforts to identify other VEEV infections.

**Figure 2.**
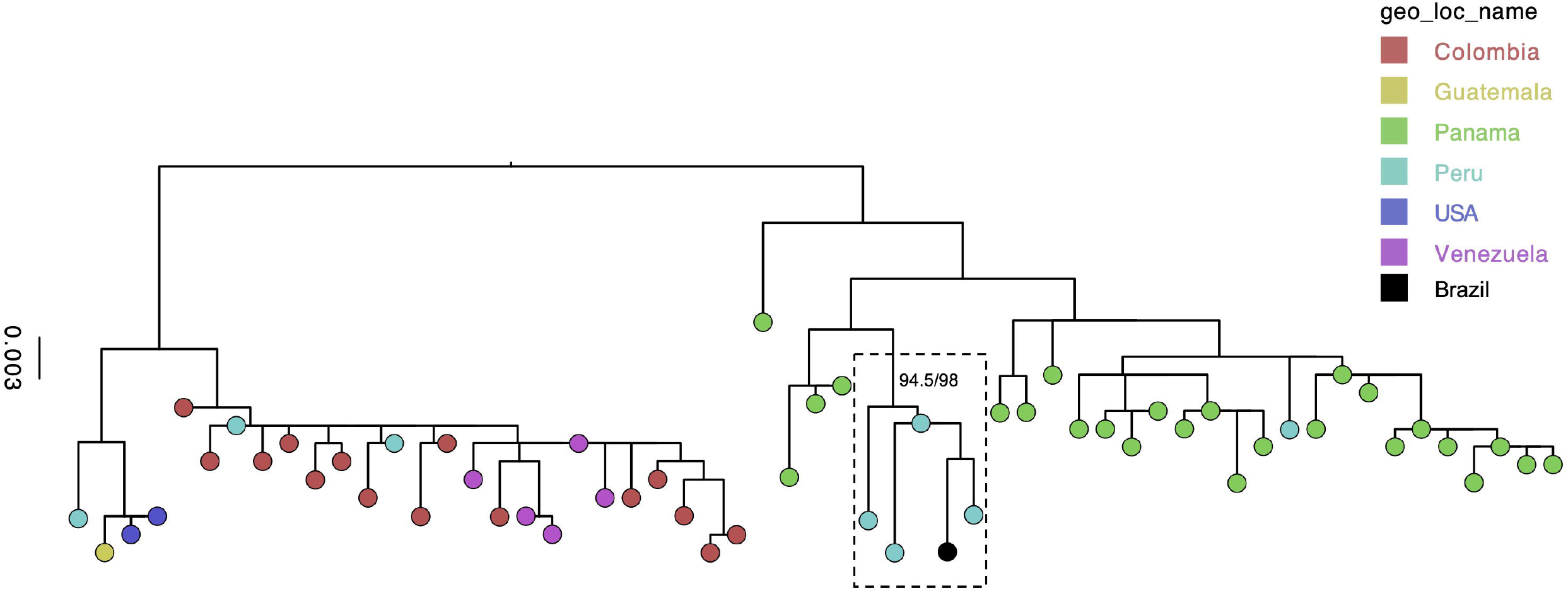
Phylogenetic analysis of the VEEV sequences. Maximum likelihood phylogenetic tree of 57 VEEV sequences related to sample Tabatinga 055 according to BLAST analysis. Scale bar indicates nucleotide substitutions per site, and tip colors represent different sampling locations (countries), as noted in the color-key boxes. The monophyletic clade containing the Tabatinga 055 sequence and Peruvian samples (all belonging to the ID genotype) is highlighted by a dashed rectangle, and the branch support (94.5/98 UFB/SH-aLRT) is shown.

## Conclusions

Despite decades of well-established evidence of Venezuelan Equine Encephalitis Complex circulation in the Brazilian Amazon, driven primarily by the enzootic strains Mucambo virus (subtype IIIA) and Pixuna virus (subtype IV) (8), no VEEV human cases have ever been officially confirmed in Brazil. Hence, we report the first laboratory-confirmed acute human VEEV infections recognized by the Brazilian National Health Surveillance System.

The molecular identification of VEEV subtype ID in Tabatinga represents a significant public health milestone in the Western Brazilian Amazon. These findings reveal VEEV as an underrecognized etiology of AFI in the region, where its nonspecific clinical presentation closely resembles endemic arboviruses such as dengue, potentially shadowing its true prevalence (2,3). This diagnostic overlap contributes to the substantial burden of undifferentiated fevers throughout the Amazon, potentially hiding the true extent of the VEEV circulation (9). Laboratory surveillance should not underestimate VEEV as an etiology of AFI in this region, where sylvatic reservoirs may sustain a continuous spillover risk to humans (10,11).

The detection of VEEV in Tabatinga is particularly alarming, given the well-documented endemic circulation and human outbreaks in neighboring countries (9,11). The phylogenetic clustering of this VEEV subtype ID strain with sequences originating from Peru (12) provides compelling molecular evidence for cross-border pathogen circulation in the Amazonian tri-border region (Brasil-Colombia-Peru). Moreover, the emergence of this enzootic strain, typically associated with sylvatic spillover from mammals, within an urban environment with competent and adaptable vectors, raises questions about local transmission dynamics and highlights a critical risk for the potential establishment of urban transmission cycles. This underscores the urgent need for enhanced surveillance and integrated clinical, reservoir, and entomological investigations in the area (10,11,13).

Finally, our results reinforce the urgent need to enhance local diagnostic capabilities by transferring critical technologies such as the VEEV real-time RT-PCR assay to frontier laboratories in the Amazon. Minimizing the detection-to-alert timeline is essential for facilitating a coordinated regional response and for implementing an effective, adaptable, and integrated disease surveillance system to address the unique challenges posed by complex and isolated border ecosystems (4,13).

The partial genome obtained in this study is available in GenBank under accession number PX115228.

## Data Availability

The partial genome obtained in this study is available in GenBank under accession number PX115228.

## Acknowledgments

We thank the study participants, the local healthcare teams in Tabatinga, the Municipal Health Secretariat of Tabatinga, and the Amazonas Health Surveillance Foundation - Dr. Rosemary Costa Pinto (FVS-RCP), for their invaluable support.

## About the Author

Dr. Santana Filho is an infectious disease medical doctor, the clinical coordinator of the Front Fever project, and a PhD. Student in the Doctoral Program in Public Health and Environment at the Sérgio Arouca National School of Public Health (ENSP), Fiocruz Rio de Janeiro, through the VigiFronteiras-Brasil (Health Surveillance in Borders) Program. His research focuses on the epidemiology of acute febrile illnesses and zoonotic disease surveillance in the Amazon border regions.

## Author contributions

F.S.S.F., D.S.V.D., G.B., M.C.S.M., and F.G.N. conceived and designed the study and contributed to data analysis. The local clinical team (FSSF, M.P.S.L.S., G.P.H.M., E.A.S.B.F., L.C.B.O., K.E.L.S., J.E.S.M., K.E.L.S., S.H.A.N., B.O.S., and K.L.O.) was responsible for patient recruitment, initial screening, case management and follow-up. M.F.S., V.C.S., V.A.N., D.S.C., T.M.O.T., M.M., F.N., D.S., M.J.P.B., and A.B.V.M. contributed to laboratory diagnostics. F.S.S.F., K.E.L.S., J.E.S.M., E.A.S.B.F., L.C.B.O., D.S.C., T.M.O.T., A.C.M.S., S.H.A.N., H.A.P.D., T.A.P.A., M.A.A.O., L.F.G., T.C.A.R., F.E.M.E., A.R.S.P, and K.E.L.S. contributed to patients, and public health surveillance data analysis. V.A.N., D.S.V.D., M.C.S.M., and F.G.N. contributed to laboratory management and obtaining financial support. F.S.S.F., G.B., M.C.S.M, and F.G.N. wrote the first draft, and all authors contributed and approved the final manuscript.

