## Supplemental Figure for "Venezuelan Equine Encephalitis Virus, Amazonas, Brazil, 2025"

**Supplemental file of the manuscript: Venezuelan Equine Encephalitis Virus, Amazonas, Brazil, 2025.**

### Methods

We used a previously designed (2017) real-time RT-PCR protocol targeting the NSP1 gene of the Venezuelan Equine Encephalitis Virus (VEEV) (Naveca, unpublished. **Table 1**), together with Taqman Fast 1-step master mix (Thermo Fisher Scientific) to detect the three cases described in this study (Cts: 35.7, 36.5, and 38.4) (**Supplemental Figure 1A**). To further confirm the results obtained using real-time PCR, we submitted the three RNA samples to two consecutive PCR amplification rounds. Initially, cDNA was produced with SuperScript IV (Thermo Fisher) and random primers. Thus, the first PCR round consisted of 35 cycles, and the second (reamplification) round consisted of 25 cycles, using a previously described pan-Alphavirus conventional RT-PCR protocol targeting the NSP1 region (Pfeffer, 1997), and 2X Platinum SuperFi PCR Master Mix (Thermo Fisher). Amplicon products were run and analyzed using Agilent TapeStation High Sensitivity D1000 ScreenTape®.

We identified one amplicon with the expected size in the reamplification assay of sample 055 (**Supplemental Figure 1B**), which was purified using AMPure XP Beads according to the manufacturer's recommendations. Patient 055's amplicon was then submitted to capillary sequencing using BigDye Terminator on an ABI 3500 nucleotide sequencer installed at the Fiocruz ILMD genomics platform (RPT01H). The ABI files were exported to Geneious Prime 2025.2.1 software and trimmed on both the 5' and 3' ends using a cut-off low-quality value of 5%. They were then used for contig assembly with the NC_001449 RefSeq as a template. Thus, the consensus sequence recovered from the forward and reverse ABI files was submitted to a BLAST search against the entire nr collection using the MegaBlast algorithm, which returned VEEV in all results with the highest score (down to 9E-157).

We then designed additional primers to amplify a larger fragment of the VEEV genome for use in phylogenetic reconstruction. This second RT-PCR amplification was conducted similarly to the first one, using random cDNA (SuperScript IV) and two consecutive PCR amplification rounds (35 cycles each) with the same primers, VEEV_Types_1PCR_FNF and VEEV_Types_FNR (1 μM each), designed in this study, and 2X Platinum SuperFi PCR Master Mix (Thermo Fisher) (**Supplemental Figure 1C**). A temperature of 52 °C was used in the hybridization step.

**Table 1. Oligonucleotides used in this study.**

| Oligo | 5'- 3' sequence | *Position | Application |
|---|---|---|---|
| VEEV_FNF | GTAGAAGCCAAGCAGGTCAC | 123-142 | Real-time RT-PCR** |
| VEEV_FNP | GACCATGCTAAYGCCAGAGC | 150-169 | |
| VEEV_FNR | GTCCACCTCCGTYTCGATCA | 212-193 | |
| VEEV_Types_1PCR_FNF | TGGAGAARGTTCACGTTGAYAT | 46-67 | External primers for nucleotide sequencing |
| VEEV_Types_FNR | GTACCCGTCGCARCTAACTATA | 899-878 | |
| VEEV_S1_2PCR_FNF | ATCCTTGACATTGGAAGTGCG | 225-245 | Internal primers for nucleotide sequencing |
| VEEV_S1_2PCR_FNR | TGGTGTCAAAGCCTATCCAGTA | 585-564 | |

* Nucleotide position related to the VEEV NC_001449 GenBank RefSeq. ** Unpublished Real-time RT-PCR protocol designed by Felipe Naveca in 2017.

The consensus 807bp sequence was then used to BLAST search against the entire nr database. All 100 hits were downloaded as a dataset that was curated for artificial sequences or sequences without complete metadata for "geographic regions of collection" or "collection data". Thus, 57 sequences were aligned with patient 055's consensus sequence using MAFFT v7.490 (1), embedded in Geneious Prime 2025.2.1. Subsequently, this file was then submitted to maximum likelihood (ML) phylogenetic inference using IQ-TREE multicore version 2.1.1 COVID-edition for Mac OS X 64-bit built Aug 20 2020 (2), with the best nucleotide substitution model selected by ModelFinder (3), and 2,000 replicates for both Ultrafast Bootstrap (4) and SH-aLRT branch support testing. The ML tree file was then edited with FigTree v1.4.4 (http://tree.bio.ed.ac.uk/software/figtree/).

**Supplemental Figure 1**

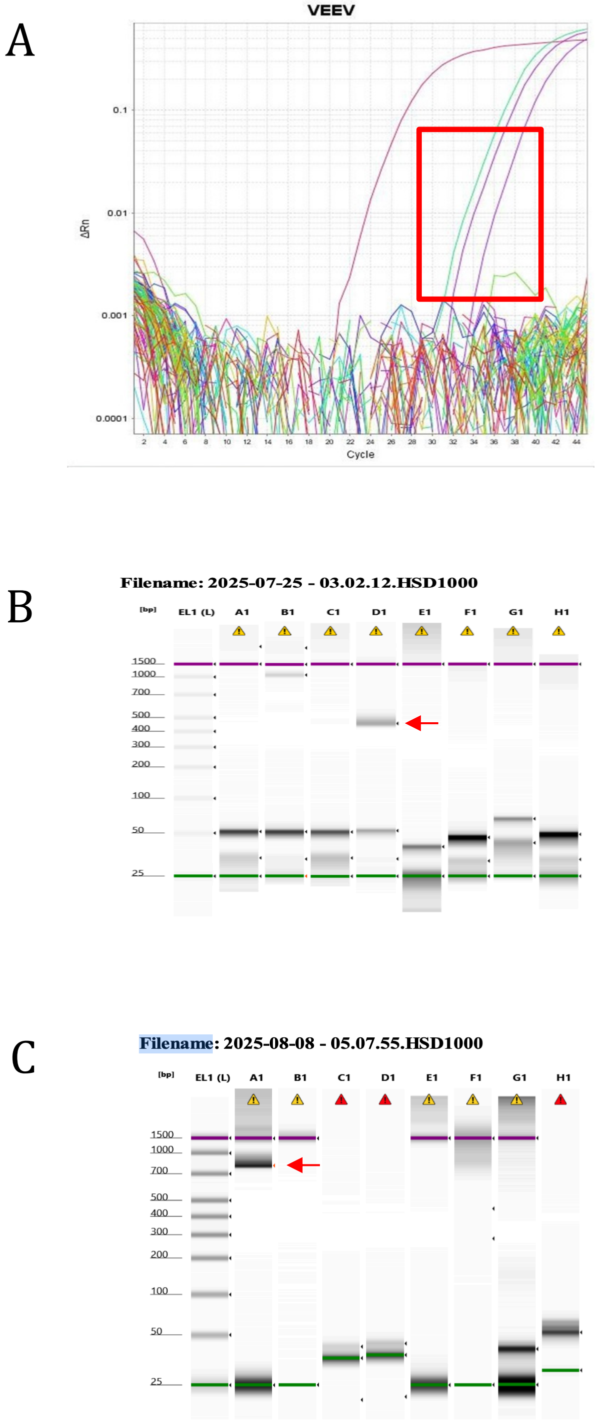


A. Amplification plot showing VEEV positive curves highlighted in red. B. RT-PCR amplification with pan-alphavirus primers. C. RT-PCR amplification with VEEV_Types_1PCR_FNF and VEEV_Types_FNR. In both B and C red arrows indicates fragments with the expected size. Other lanes contain samples not amplified pan-alpha (B) or other evaluated temperature or primer concentrations (C).

**Supplemental file references**


1. Katoh K, Standley DM. MAFFT multiple sequence alignment software version 7: improvements in performance and usability. Mol Biol Evol. 2013 Apr;30(4):772-80. doi: 10.1093/molbev/mst010.
2. Minh BQ, Schmidt HA, Chernomor O, Schrempf D, Woodhams MD, von Haeseler A, Lanfear R. IQ-TREE 2: New Models and Efficient Methods for Phylogenetic Inference in the Genomic Era. Mol Biol Evol. 2020 May 1;37(5):1530-1534. doi: 10.1093/molbev/msaa015. Erratum in: Mol Biol Evol. 2020 Aug 1;37(8):2461. doi: 10.1093/molbev/msaa131.
3. Kalyaanamoorthy S, Minh BQ, Wong TKF, von Haeseler A, Jermiin LS. ModelFinder: fast model selection for accurate phylogenetic estimates. Nat Methods. 2017 Jun;14(6):587-589. doi: 10.1038/nmeth.4285.
4. Hoang DT, Chernomor O, von Haeseler A, Minh BQ, Vinh LS. UFBoot2: Improving the Ultrafast Bootstrap Approximation. Mol Biol Evol. 2018 Feb 1;35(2):518-522. doi: 10.1093/molbev/msx281.
